# Predicting the long-term impact of rotavirus vaccination in 112 countries from 2006-2034: a transmission modeling analysis

**DOI:** 10.1101/2022.09.23.22280291

**Authors:** A.N.M. Kraay, M.K. Steele, J.M. Baker, E.W. Hall, A. Deshpande, B.F. Saidzosa, A. Mukaratirwa, A. Boula, E. M. Mpabalwani, N.M. Kiulia, E Tsolenyanu, C. Enweronu-Laryea, A. Abebe, B. Beyene, M. Tefera, R. Willilo, N. Batmunkh, R. Pastore, J.M. Mwenda, S. Antoni, A.L. Cohen, V.E. Pitzer, B.A. Lopman

## Abstract

Rotavirus vaccination has been shown to reduce rotavirus burden in many countries, but the long-term magnitude of vaccine impacts is unclear, particularly in low-income countries. We use a transmission model to estimate the long-term impact of rotavirus vaccination on deaths and disability adjusted life years (DALYs) from 2006-2034 for 112 low- and middle-income countries. We also explore the predicted effectiveness of a one- vs two-dose series and the relative contribution of direct vs indirect effects to overall impacts. To validate the model, we compare predicted percent reductions in severe rotavirus cases with the percent reduction in rotavirus positivity among gastroenteritis hospital admissions for 10 countries with pre- and post-vaccine introduction data. We estimate that vaccination would reduce deaths from rotavirus by 49.1% (95% UI: 46.6–54.3%) by 2034 under realistic coverage scenarios, compared to a scenario without vaccination. Most of this benefit is due to direct benefit to vaccinated individuals (explaining 69-97% of the overall impact), but indirect protection also appears to enhance impacts. We find that a one-dose schedule would only be about 57% as effective as a two-dose schedule 12 years after vaccine introduction. Our model closely reproduced observed reductions in rotavirus positivity in the first few years after vaccine introduction in select countries. Rotavirus vaccination is likely to have a substantial impact on rotavirus gastroenteritis and its mortality burden. To sustain this benefit, the complete series of doses is needed.

---

Diarrhea is a leading cause of under-five mortality worldwide (1), and rotavirus is a predominant etiology of diarrhea mortality (2), estimated to cause 128,500 deaths worldwide as of 2016 (3). In 2009, two live, attenuated oral rotavirus vaccines were recommended for global use: Rotarix, a monovalent vaccine administered in a two-dose series, and RotaTeq, a pentavalent vaccine with a three-dose series. In recent years, two additional three-dose vaccines have become available, Rotavac and ROTASIIL, which are also live oral vaccines with similar safety and efficacy profiles. All four vaccines have received WHO prequalification (4), and Rotarix is currently the product most used in low- and middle-income countries that have introduced rotavirus vaccination into their national immunization programs (5). While vaccine efficacy is high in high-income countries (84 to 90%), there is a gradient of lower protection in middle- (∼75%) and lower-income settings (∼50%) where burden is highest (6).

While some early-introducing countries integrated rotavirus vaccine into their national immunization programs as early as 2006, many African nations did not implement rotavirus vaccination until 2012 or later (5), and most low- and middle-income countries (LMICs) in Asia have not yet introduced rotavirus vaccines. The long-term impacts of rotavirus vaccination, particularly in LMICs, are unclear and will require further evaluation. Long-term transmission dynamics may differ from those in the first few years after licensure based on the steady-state levels of protection among children overall, due to gradually increasing vaccine coverage among young children, aging of the initial cohort of vaccinated individuals and complex dynamics of indirect benefits (7).

The real-world impact of rotavirus vaccination will depend on several factors, including direct efficacy in a specific population, the completeness of vaccination (with respect to both coverage and number of doses), changing demographics, and indirect impacts for unvaccinated individuals. Although a full 2- or 3-dose course is recommended, some children do not complete the schedule. In observational studies, a recent meta-analysis showed that the average difference in efficacy of one dose of Rotarix is about 19% lower than the full two-dose series, with the largest differences observed in settings with high child mortality and low income (6).

In high-income settings, indirect effects have been observed, whereby unvaccinated individuals also had lower risk of hospitalization for rotavirus following vaccine introduction (8). Across several middle- and high-income countries, the overall reduction in severe rotavirus attributable to indirect effects has been estimated as 22% (9). It remains unclear if these benefits will be present in low-income countries, where vaccine efficacy is generally lower (10) and the force of infection is higher (11). The potential strength of indirect effects might increase or decrease over time depending on underlying transmission and immunity patterns. Higher vaccine coverage achieved as rollout proceeds could enhance indirect protection, but the impact could also decrease over time as the number of susceptible individuals accrued eventually becomes sufficient to sustain new waves of transmission.

In this analysis, we estimate the potential impact of rotavirus vaccination in 112 countries from 2006 to 2034. For four large countries (Pakistan, India, Nigeria, and Ethiopia, hereafter called PINE countries), we also predict the potential magnitude of indirect effects for and examine how a one-dose schedule might influence overall population impact. These four countries have high rotavirus disease burden that have been prioritized for increasing vaccine uptake by both Gavi and the Gates Foundation. All simulations were done as part of the Gavi-supported Vaccine Impact Modeling Consortium (VIMC), which aims to produce robust estimates of vaccine impact for ten vaccine-preventable diseases to help guide future vaccine rollout decisions (12).

## Methods

Our deterministic, age-structured compartmental model simulates rotavirus transmission and estimates disease incidence/burden by country. The model is based on a Susceptible– Infected–Recovered (SIR) structure, with elaborations to capture incremental acquisition of immunity and transmission of rotavirus (Figure 1). We model the following age groups: 0-1 months, 2-3 months, 4-11 months, 1-year age bands from 1 to 4 years old, and 5 years and older, similar to other published rotavirus transmission models (reviewed in (13)). We use realistic country-specific, age-specific population sizes, aging and death rates (14). All main text results focus on children under 5 years of age.

**Figure 1.**
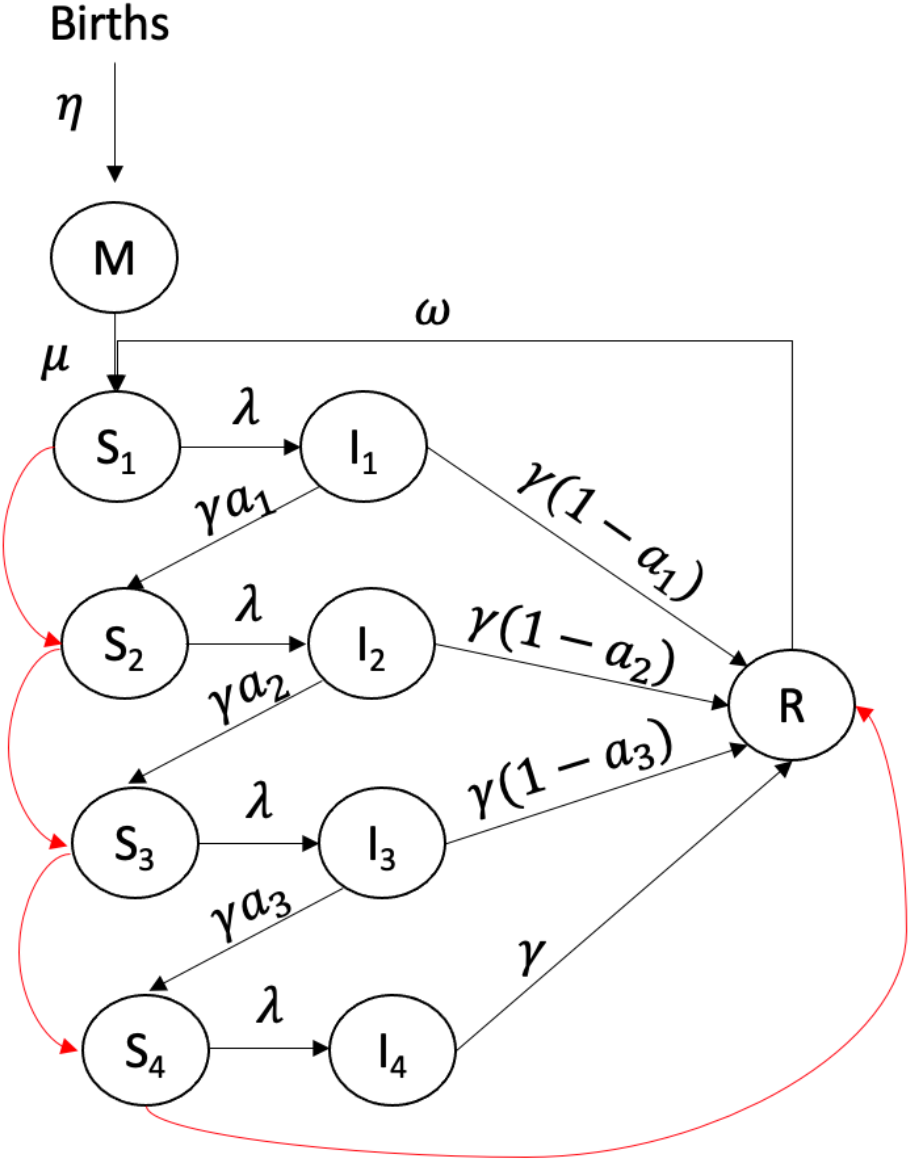
Model structure. Vaccination is indicated by the red arrows. We also separately tracked vaccinated and unvaccinated populations. Children are born with maternal immunity (M class). After this immunity wanes, children are fully susceptible to rotavirus infection (entering the S_1_ class). Upon infection, children become infectious (entering the I class), after which they may either develop long term immunity (entering the R class) or enter a lower susceptibility class (progressing to the next susceptible compartment, S). Secondary, tertiary, and quaternary infections are less likely to result in severe disease or death. After four infections, children enter the long-term immunity class (R). Long-term immunity wanes over time, after which children are fully susceptible to subsequent rotavirus infections (entering the S_1_ class). Vaccination is assumed to act like one natural infection, and vaccinated individuals who respond to the vaccine move up one infection class after each dose. Model parameters and equations are shown in the appendix.

Infants are born into the model with maternal immunity (15). After maternal immunity wanes, infants become susceptible to a primary rotavirus infection. We assume protection is conferred by previous infections against subsequent infections, such that the proportion of individuals that remain susceptible to re-infection decreases with each subsequent infection (16). All infections are assumed to have the same duration of infectiousness; however, non-primary infections have lower per-contact infectiousness relative to primary infections (13). Immunity is assumed to be “take-type”, whereby a portion of individuals develop long-term immunity while others remain fully susceptible to subsequent infection. We assume primary, secondary, tertiary, and quaternary infections had different probabilities for developing rotavirus gastroenteritis (GE) as per (16). We assume only severe rotavirus GE cases are reported to surveillance and can result in death.

This model incorporates the introduction of vaccines in a specified year, delivered to 2- and 4-month-olds (Rotarix-like). We incorporate an immunogenicity parameter that determines whether individuals respond to the vaccine, which is lowest in low-income countries (Table A1). If individuals respond to vaccination, then we assume that vaccine immunity acts like a natural infection (13); the probability of becoming infected decreases with each subsequent vaccine dose and natural infection (Figure 1). Values for natural history parameters were set to values identified in birth cohort and challenge studies (Table A1). Specifically, parameters related to severity of primary and subsequent infections were set based on a birth cohort study from India (17) and parameters related to the duration of immunity and length of infection were set based on data from Brazil (15).

We used a linear regression model to estimate the mean age of severe rotavirus infection, and subsequently calculated the basic reproduction number (R_0_) (described below). We initially aimed to fit the model to estimate country-specific effective contact rates, but this approach was computationally intensive and was less accurate than the regression approach. We therefore focus on results using the regression approach here.

### Estimate of R_0_ by regression model of mean age of severe disease

We calculated R_0_ from the mean age of infection as follows:

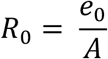

where *e*_*0*_ represents the country-specific life expectancy at birth (in years) and *A* is the mean age of severe infection (in years) estimated from linear regression (described below). The value of *e*_*0*_ is based on the 2017 estimates from United Nations World Population Prospect (14).

We used a linear regression model to estimate the mean age of severe rotavirus GE for 112 countries. The regression model was fit to surveillance data from 112 countries in the Global Rotavirus Surveillance Network (GRSN)(18). For each country, we calculated the average age in weeks of children under five with medically attended (clinic visit or hospital admission) rotavirus diarrhea. While the overall number is the same, the countries included for regression model fitting included some countries for which vaccine impacts were not modeled, and not all countries included in the model analysis had GRSN data. The final model was selected based on fit statistics and prediction accuracy, and included: under 5 mortality rate, birth rate, life expectancy, percent of population living in a rural setting, and total gross domestic product. See appendix for regression model details and performance (Table A2, Figure A1). We applied the same regression model to countries lacking surveillance data.

### Uncertainty analysis

Model simulation, fitting, and analysis were conducted in *R* version 3.4.0 using the *deSolve* package (19). For probabilistic model runs, we generated 200 parameter sets by uniformly sampling from the published range of vaccine immunogenicity and the 95% confidence interval of the estimated mean age of infection from the regression model (see Appendix). We then simulated the model for each set of fixed and sampled parameters. We calculated the central burden/impact estimate as the median of the 200 probabilistic runs.

### Estimates of vaccine impact

We predicted the number of deaths and DALYs attributable to rotavirus infections among children under 5 years old expected in all 112 countries under three scenarios: no vaccination, a default vaccination scenario, and a best-case vaccination scenario. Coverage estimates for both vaccination scenarios were provided by the Vaccine Impact Modelling Consortium and were based on feasible country-specific coverage levels either with (best case) or without (default) additional vaccine campaigns (20) (see Appendix). To estimate the impact of vaccination on deaths and DALYs averted, we compared all vaccine impacts with a no vaccination scenario (see Appendix). For PINE countries, we examined the difference between direct and overall vaccine effects and considered the degree to which vaccine impacts might be affected if a one-dose vaccine schedule were used. For both analyses, we used the default vaccine coverage estimates.

To assess the importance of indirect effects, we first allowed the force of infection to change over time as vaccination was introduced, yielding an estimate of the overall effect of vaccination. Second, we fixed the force of infection for these four countries to its value just before vaccination was introduced (21), yielding an estimate of the direct effect of vaccination. The indirect effect of vaccination can then be estimated as the overall effect minus the direct effect (9).

For one-dose vaccine simulations, we assumed that that the probability of receiving first and second doses are independent so that the fraction of the population receiving only one dose was the square root of estimated proportion of the population receiving two doses (i.e., when two-dose vaccine coverage was 30%, one-dose coverage would be 55%). We also assumed that individuals receiving only one dose of vaccine received this dose at 2 months of age.

### Model validation

Model predictions for severe rotavirus cases were compared with data on rotavirus positivity among GE hospitalizations from 10 countries from GRSN (18): eight from Africa (Cameroon, Ethiopia, Ghana, Kenya, Tanzania, Togo, Zambia, and Zimbabwe), one from Europe (Tajikistan), and one from the western Pacific region (Fiji). For the countries included in the analysis, the overall number of GE hospitalizations and rotavirus positive hospitalizations varied widely in the years prior to vaccine introduction due to a variety of external factors, but the percent positivity was more stable (Table A3). Prior work has shown that the percent positivity and overall rotavirus positive hospital admissions have similar trends when annual tests are stable (22). Thus, we validated our model using reductions in percent positivity but also considered model consistency with counts of rotavirus positive hospital admissions as a sensitivity analysis (Figures A2 and A3).

To calculate the percent rotavirus positivity, we divided the number of cases with gastrointestinal symptoms admitted to the hospital who were positive for rotavirus by the total number of cases with gastrointestinal symptoms admitted to the hospital who were either positive or negative for rotavirus. We estimated the percent reduction in severe rotavirus cases for each year since vaccination from our transmission model, compared with the pre-vaccine period. The number of severe rotavirus cases in the pre-vaccine period was averaged across all years of available pre-vaccine data for each country. For each comparison, we calculated the 95% confidence interval for the empirical GRSN data and the 95% uncertainty intervals (UI) across 200 probabilistic runs for the modeled impacts. For UIs, the lower bound was the 2.5^th^ percentile of the distribution and the upper bound was the 97.5^th^ percentile of the distribution. See Table A3 and accompanying text for more details.

## Results

### Overall impacts across all 112 countries

Overall, our models predicted that a total of 102,000 (95% UI: 96,000–112,000) deaths per year would be averted among children under five for all 112 countries considered by 2034 under the default scenario, and 110,000 (95% UI: 103,000–121,000) deaths per year would be averted under the best-case scenario. These values correspond to a 49.1% (95% UI: 46.6–54.3%) decline in annual deaths for the default scenario in 2034 or 52.7% (95% UI: 50.0–58.6%) for the best-case scenario compared with no vaccination. A total of 1.59 million (95% UI: 1.50 – 1.75 million) deaths could be averted between 2006 and 2034 in the best-case scenario compared to the no vaccination scenario. Regionally, the total number of yearly deaths averted by 2034 was expected to be greatest in sub-Saharan Africa (77,000 (95% UI: 72,000– 85,000) to 83,000 (95% UI: 77,000 –92,000) deaths averted/year, a 47.7% (95% UI: 45.0—53.3%) to 51.4% (95% UI: 48.4 – 57.7%) reduction), followed by South Asia (18,000 (95% UI: 16,000–21,000) to 18,000 (95% UI: 16,000–21,000) deaths averted/year, a 56.8% (95% UI: 49.6—64.8%) to 57.5% (95% UI: 50.2–65.5%) reduction) (Figure 2). Vaccination was expected to reduce deaths most in lower-middle-income countries, corresponding to the regions with highest death burden due to rotavirus. However, other regions also had substantial impacts, with reductions of 44.9% (95% UI: 39.5—49.9%) to 57.9% (95% UI: 51.9 –64.1%) in East Asia & the Pacific, 49.6% (95% UI: 45.6–55.8%) to 50.4% (95% UI: 46.4–56.7%) in Europe and Central Asia, 53.9% (95% UI: 49.3 –59.0%) to 54.9% (95% UI: 50.3 –60.1%) in Latin America & the Caribbean, and 48.8% (95% UI: 46.8 – 58.3%) to 51.9% (95% UI: 44.1–54.9%) in the Middle East & North Africa.

**Figure 2:**
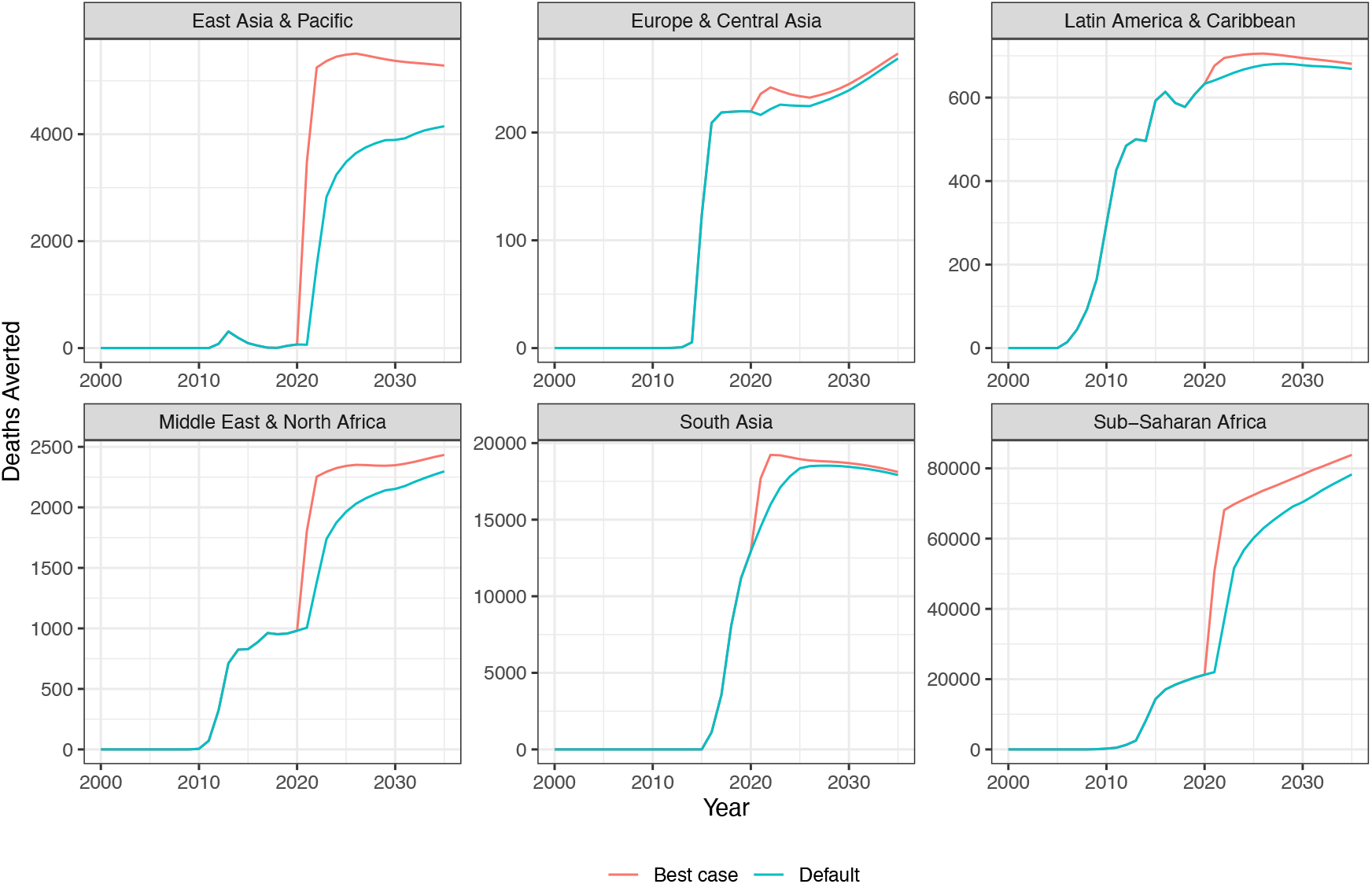
Predicted annual deaths averted by region under the default vaccination scenario. For similar figures for DALYs averted, see Figures A4 and A5.

While our model predicted slight increases in rotavirus deaths among 3- and 4-year-old children due to shifting of disease burden to older age groups (Figure 3), the net impact of vaccination was overwhelmingly beneficial for all 112 countries due to dramatic reductions in deaths among children under 3 years of age (Figure 2, Figure 3).

**Figure 3:**
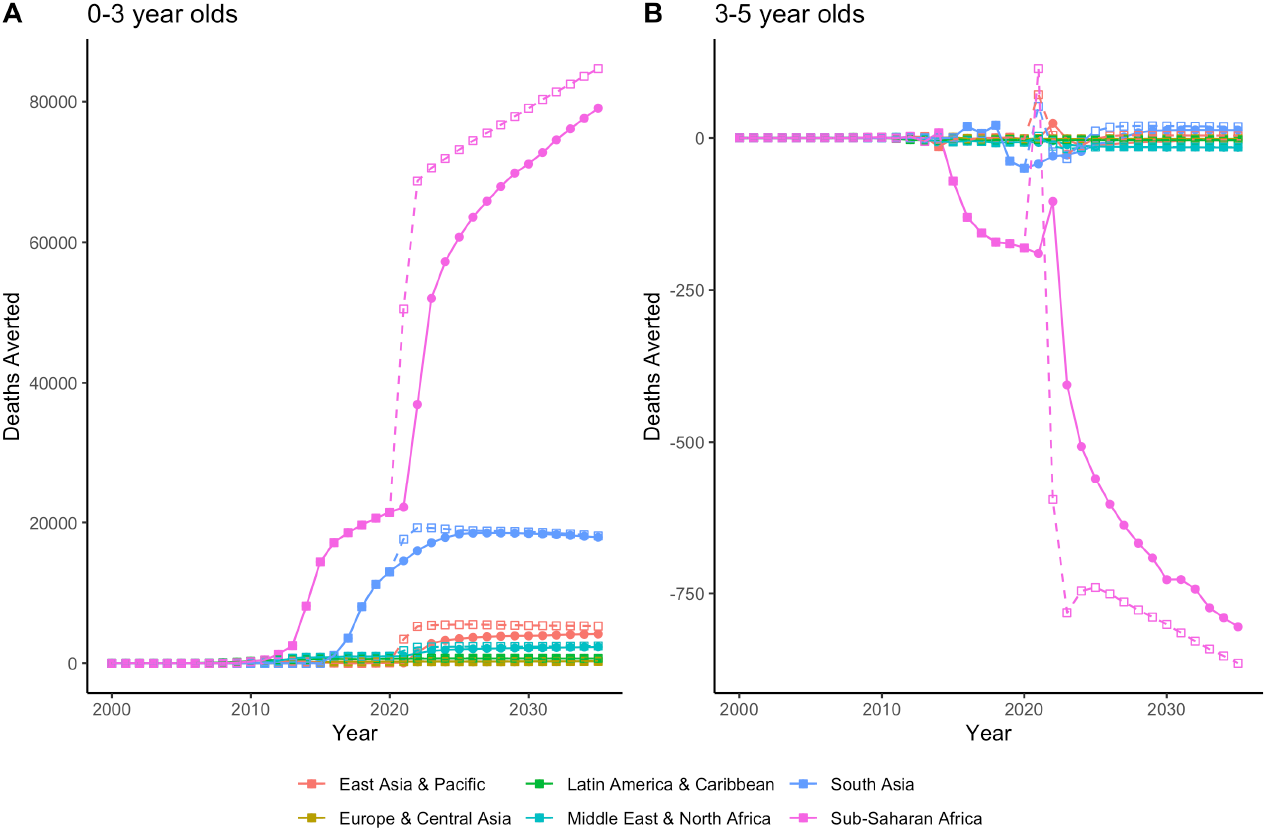
Deaths averted by age group, region, and vaccine coverage scenario over time for A) 0-3 year olds and B) 3-5 year olds. Solid lines and circles show predicted impacts for the default scenario and dashed lines and open square shapes show impacts for the best-case scenario. See Figure A6 for plots of total deaths stratified by yearly age groups.

### Additional analysis for PINE countries

Overall, vaccination was expected to avert 51.4% (UI: 46.4—59.1%) to 55.4% (UI: 50.0—64.0%) of all rotavirus deaths per year by 2034 in PINE countries. Most of this predicted benefit was due to the direct effect of rotavirus vaccination (Figure 4, Table 1), with direct effects accounting for 69-97% of the overall vaccine impact (82.3% for Pakistan (95% UI: 75.1-85.9%), 69.4% for India (95% UI: 62.7-74.9%), 97.1% for Nigeria (95% UI: 85.3-99.7%), and 81.2% for Ethiopia (95% UI: 71.5-83.7%)). Indirect effects were only observed for children under 1 year of age, after which point immunity due to natural infection became more important given the increasing fraction of individuals having had at least 1 natural infection. Indirect effects were least beneficial in Nigeria, where the birth rate is highest and average age of first infection is lowest. Indirect benefits became more pronounced over time as the overall coverage of vaccination increased.

**Table 1.** Percent of rotavirus deaths averted 12 years after vaccine introduction due to rotavirus vaccination assuming default vaccine coverage levels.

| Country | Direct Effect (DE)<br>Point Estimate<br>(95% UI) | Overall Effect (OE)<br>Point Estimate<br>(95% UI) | Indirect effect (OE-DE)<br>Point Estimate<br>(95% UI) | Fraction of benefit due to direct effect (DE/OE)<br>Point Estimate<br>(95% UI) |
| --- | --- | --- | --- | --- |
| Pakistan | 45.7%<br>(39.2—51.7%) | 55.3%<br>(48.1—67.1%) | 9.6%<br>(6.8—15.8%) | 82.3%<br>(75.1—85.9%) |
| India | 40.3%<br>(34.6—46.4%) | 58.1%<br>(49.1—67.2%) | 17.8%<br>(13.5—22.9%) | 69.4%<br>(62.7—74.9%) |
| Nigeria | 47.5%<br>(40.8—52.7%) | 48.7%<br>(42.6—58.7%) | 1.2%<br>(0.4—8.4%) | 97.1%<br>(85.3—99.7%) |
| Ethiopia | 37.5%<br>(34.6—39.5%) | 46.2%<br>(42.4—52.2%) | 8.7%<br>(7.0—14.7%) | 81.2%<br>(71.5—83.7%) |

**Figure 4.**
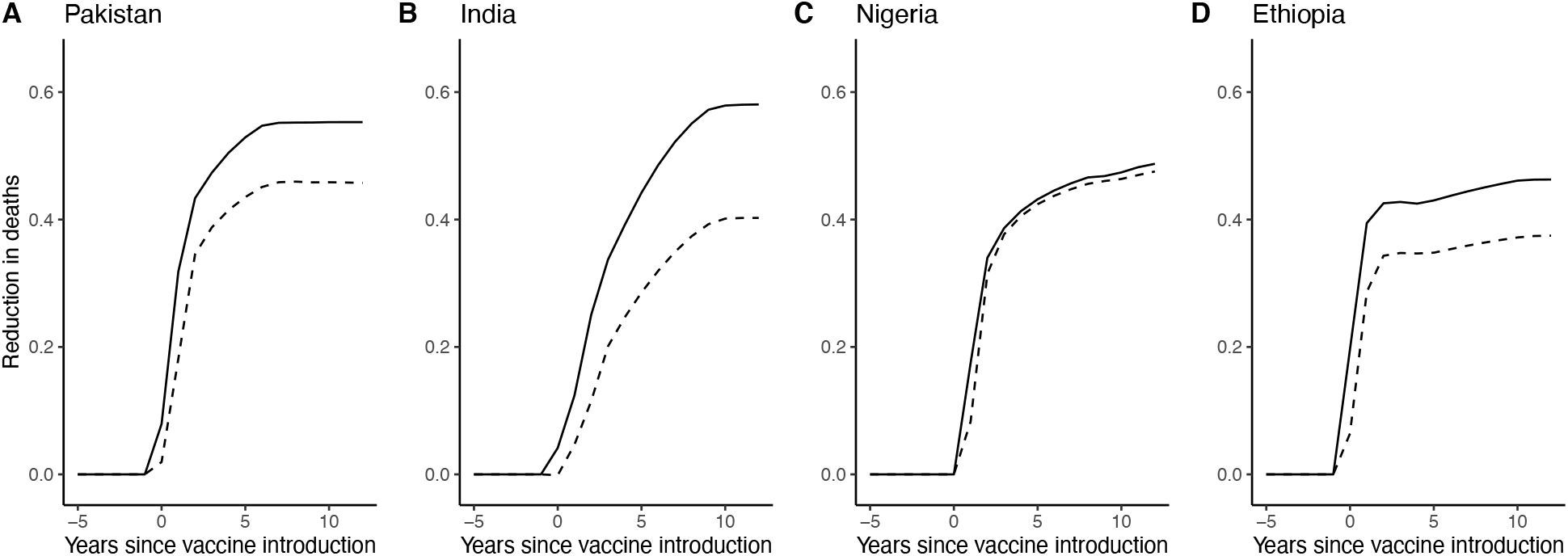
Direct vaccine effects only (dashed line) and overall vaccines effects (solid line) up to 12 years post vaccine introduction in PINE countries. Panels show A) Pakistan, B) India, C) Nigeria, and D) Ethiopia.

While a one-dose schedule was predicted to be relatively similar in performance in the first few years after vaccine introduction compared to a two-dose schedule, the one-dose schedule was ultimately about half as impactful as the two-dose series after coverage among children under five stabilized to its steady state values (Figure 5). For example, the first year after vaccine introduction, a one-dose schedule was 52-80% as impactful as a two-dose schedule. However, at 4 years after introduction, the one-dose schedule was 58-65% as impactful as a two-dose schedule and the relative benefit between the two appeared to have mostly stabilized. For all four countries, a one-dose schedule was predicted to ultimately perform 57-63% as well as two doses after 12 years (Pakistan: 57.7%, 95% UI: 56.0-59.0%; India: 56.7%, 95% UI: 55.6-58.9%; Nigeria: 62.5%, 95% UI: 58.7-62.7%; Ethiopia: 56.8%, 95% UI: 55.4-57.6%). In India and Pakistan, outcomes were initially more similar for a one-compared with two-dose schedule, likely because initial vaccine coverage was much lower in these two countries.

**Figure 5.**
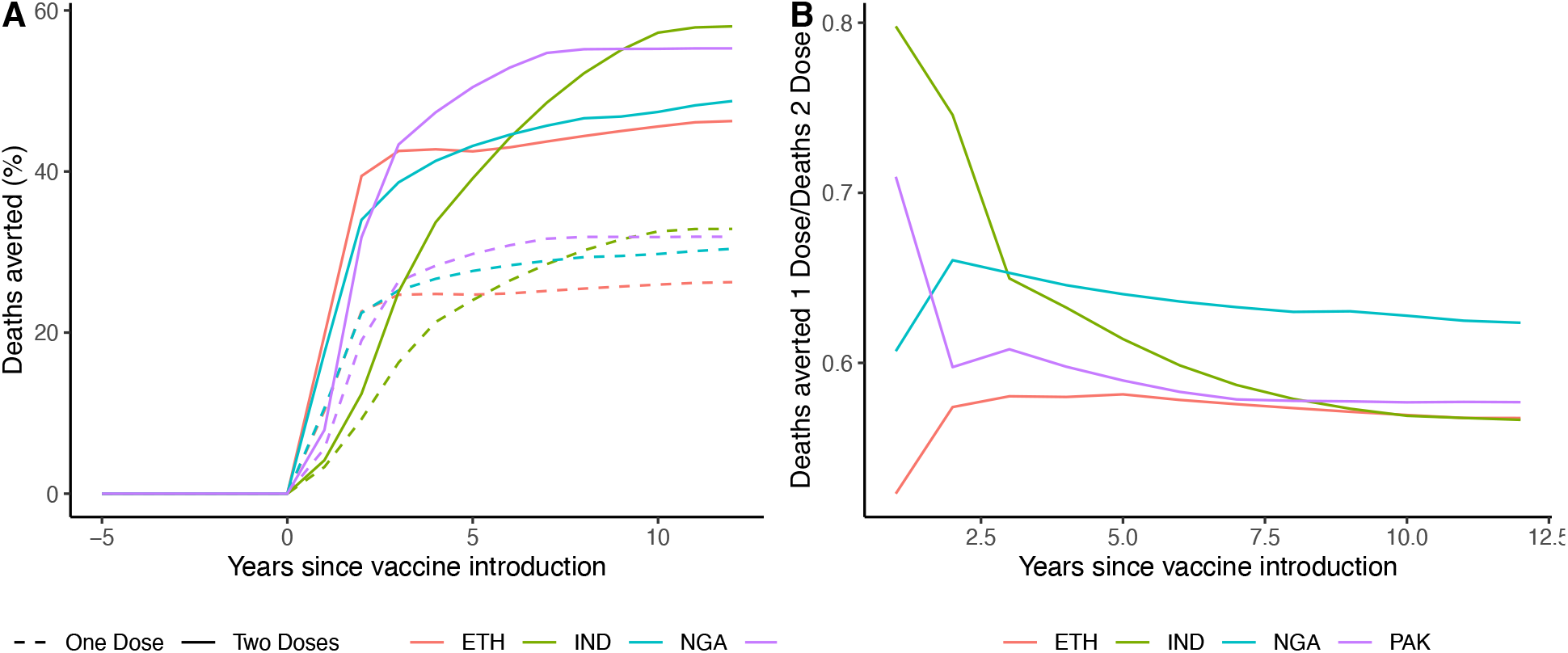
A comparison of the impact of a one- vs. two-dose vaccine schedule for the first 12 years after vaccine introduction. A) The percent of deaths averted, and B) the relative performance of a one vs. two dose schedule.

### Validation with GRSN data

For most countries, the 95% confidence intervals for reductions in rotavirus hospitalizations within GRSN overlapped the 95% uncertainty interval for the relative decrease in severe rotavirus cases produced by our transmission model (Fig 6, Fig 7). For children under 1 year of age, among whom rotavirus is more likely to be severe, the confidence intervals were overlapping for almost all countries and years except Zambia. When compared with rotavirus positive admissions (unscaled by total GE hospitalization admissions) findings were similar, but there was a greater discrepancy between modeled and observed cases (Figure A2, Figure A3).

**Figure 6:**
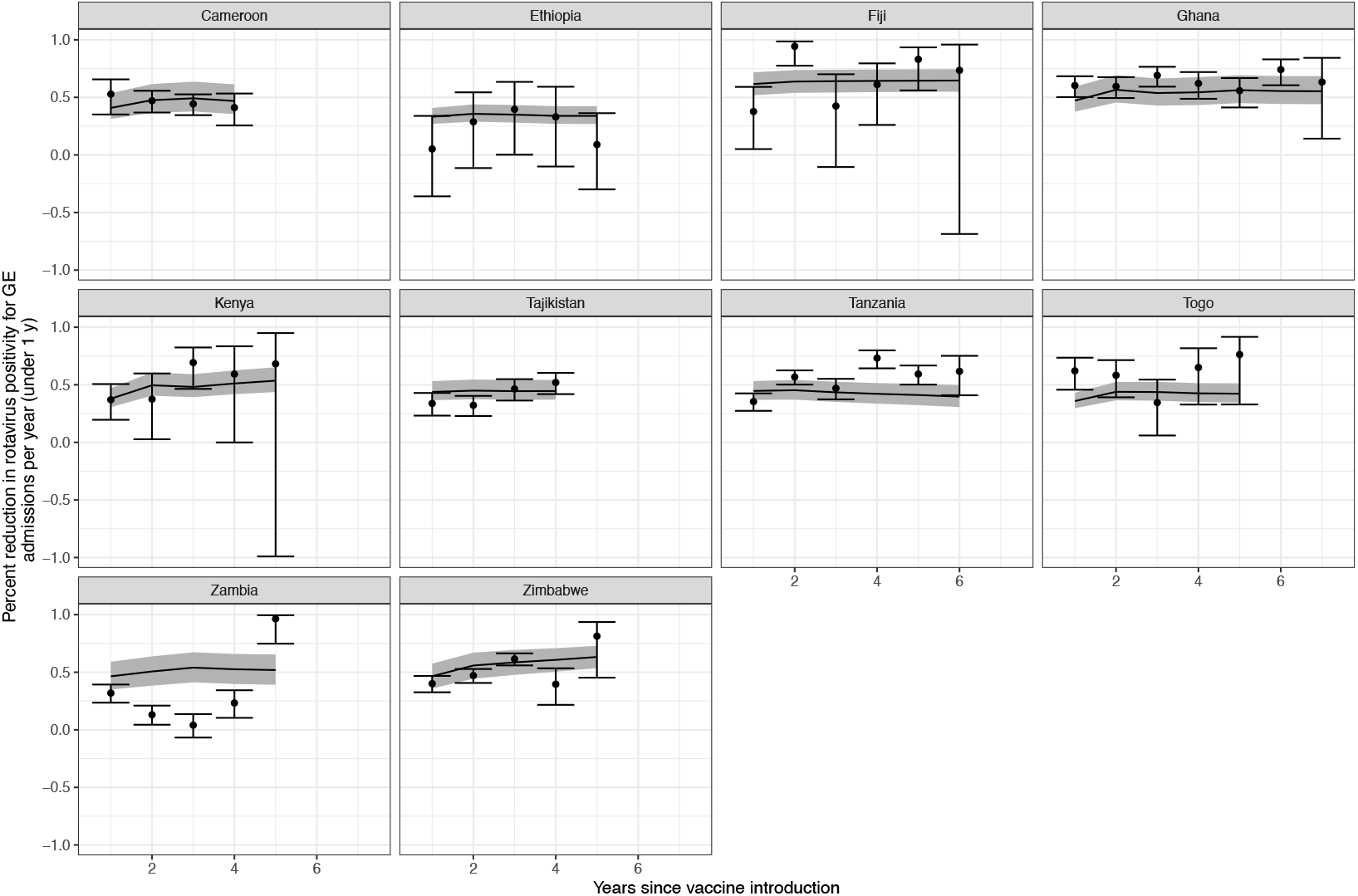
Predicted vaccine impacts on annual severe rotavirus cases for children under 1 year of age (model) and percent reduction in rotavirus positivity (GRSN data) (y-axis) by number of years since vaccine introduction (x-axis) for 10 countries (shown in different panels). Modeled impacts are shown in solid lines, with the grey ribbon showing 95% simulation intervals. Impacts based on surveillance data are shown with points, with error bars to show uncertainty (error bars were calculated using standard methods, assuming that the percent reduction in positivity approximated a risk ratio).

**Figure 7.**
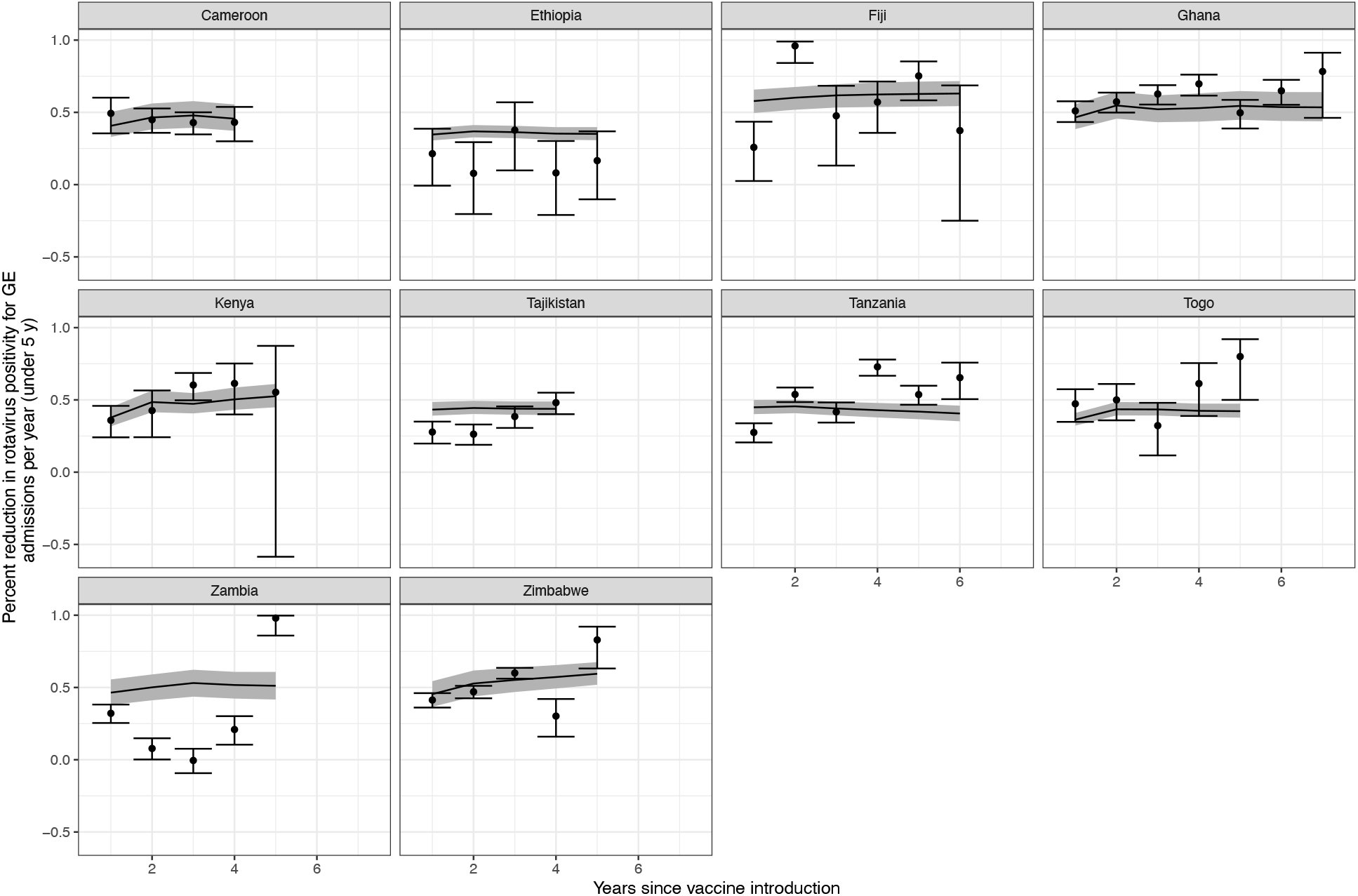
Predicted vaccine impacts on annual severe rotavirus cases for children under 5 years of age (model) and percent reduction in rotavirus positivity (GRSN data) (y-axis) by number of years since vaccine introduction (x-axis) for 10 countries (shown in different panels). Modeled impacts are shown in solid lines, with the grey ribbon showing 95% credible intervals. Impacts based on GRSN surveillance data are shown with points, with error bars to show uncertainty (error bars were calculated using standard methods, assuming that the percent reduction in positivity approximated a risk ratio).

## Discussion

Our model predicts a strong and sustained impact of rotavirus vaccination worldwide. Overall, we predict that 105,000 (95% UI: 99,000 – 115,000) deaths could be averted per year, representing a reduction of 50.4% of rotavirus deaths (95% UI: 47.4%–55.2%) compared to a scenario with no vaccination. While additional efforts to increase coverage (the best-case scenario) did provide additional benefit compared to the default scenario, this increase was modest, suggesting that substantial benefits can be achieved in many countries even under routine immunization practices. The rollout of vaccination is also predicted to shift disease burden to older age groups, particularly 3- and 4-year-old children. The absolute impact in terms of deaths and DALYs averted by vaccination is predicted to be greatest in countries with the greatest disease burden, despite lower vaccine performance. While most of the benefit is attributable to the direct effect of vaccination, indirect effects also increase the overall impact several years after vaccine introduction when coverage reaches substantial levels. While administering only one dose of vaccine provides protection, in the long-term, using a one-dose schedule is only about half as effective as a full two-dose schedule.

The consistency between our modeled impacts and observed reductions in rotavirus positivity in the first few years after introduction adds confidence to our longer-term projections. Small differences might be explained by several factors. First, due to the lack of reliable natural history data from different countries, we used data from a single study in India to parameterize our transmission model, which suggests that natural rotavirus infection confers immunity against reinfection, but provides little protection against progression to disease (17). This assumption might not hold uniformly in all countries if, for example, immunity against disease is stronger in middle-income settings (23). Moreover, vaccine immunogenicity may vary by country (24). It is also possible that vaccine performance may improve over time if the background rate of enteric infection is reduced (25) through greater access to improved water, sanitation, and hygiene (WASH) (26). Second, to validate our model, we compare predicted severe rotavirus cases with rotavirus hospitalizations, which may only be the most severe subset of severe rotavirus infections. Rotavirus vaccination is known to be most effective against more severe disease (27), so this factor could cause downward bias in our impact estimates.

In our model, indirect effects did appear to increase the overall impact of vaccination beyond what would be expected by direct effects alone, even in low-income countries. The magnitude of this benefit varied by country. The overall difference between the direct effect and overall effect ranged from 1.2%-17.8%, which is somewhat lower than has been found in middle- to high-income countries (22%) (9). Our indirect effect estimates for India are substantially higher than were estimated previously by Rose et al., who found minimal evidence for indirect effects (28). The primary reason for this difference is likely that we include a long-term immune class for both vaccinated and unvaccinated individuals, which Rose et al do not include, and from which waning is extremely slow. Given that the direct effect of vaccination is primarily related to severe disease and is experienced even for individuals who do not develop long-term immunity, the existence of this long-term immune state would increase the hypothetical magnitude of indirect effects without substantially changing the magnitude of direct effects. In our model, the strength of indirect effects appeared to be highest for countries with lower birth rates and lower overall transmission rates (approximated by a higher age of first infection). As both birth rates and background transmission rates are higher in LMICs than high-income countries, indirect effects would be expected to be weaker in these sites. More empirical data on indirect effects in low-income countries are needed to validate this prediction.

Likely in part due to the inclusion of indirect effects in this dynamic model, our estimates of deaths averted are somewhat higher than predicted by static models previously. Our model and two other static models were included in ensemble predictions for deaths averted by Toor et al for the same 112 countries included in this manuscript (20). In that analysis, 840,000 deaths were expected to be averted in the 112 countries between 2000 and 2030 when combining estimates from all models. Over the same time frame and for the same countries, our model predicted 974,000 deaths will be averted, an estimate that is about 16% higher than the combined ensemble estimate.

Due to the speed at which vaccine coverage is expected to be increased and our assumption that each dose provides similar protection to one natural infection, we found that the relative impact of one-compared with two-dose schedules was lower than has been reported previously in observational studies, but are similar to predictions from a different modeling study, which found 63% relative efficacy for 1 compared with 2 doses (29). The difference in impacts at 1 vs. 4 years post-introduction relates to the time scale at which vaccine coverage can be increased and the extent to which a one-dose schedule can increase the number of individuals vaccinated. Because we assume that vaccine coverage for a one-dose series is the square root of coverage for a two-dose series, a one-dose schedule can greatly increase vaccine coverage early in rollout; but, because one dose has lower efficacy than the full two-dose series, this benefit may not overcome reduced vaccine efficacy. Our model predicts single-dose vaccination being only about 50% as effective as a two-dose series compared with about 81% in prior studies overall, with a bigger difference in countries with higher under five mortality (reviewed in (6)). The larger relative benefit of two doses found in our study is likely partially attributable to the fact that our analysis focused on countries with medium to high under five mortality rates. Additionally, the observational studies used to produce these estimates had shorter follow-up duration than we simulated. In our analysis, the short-term benefits of a one-dose schedule are outweighed by two-dose regimen since coverage saturates at ∼90%, but vaccine performance is compromised if a second dose is not used.

Our study has several limitations. First, we assume that rotavirus vaccines are administered at exactly 2 and 4 months of age, which is similar to the dosing schedule recommended with vaccine licensure for Rotarix(27). However, this assumption may lead to an overestimate of impact in countries where rotavirus vaccines are administered later if natural infections have already occurred. The actual timeline of administration varies for individual countries due to alternative vaccine schedules (30) or late receipt of vaccine, which is especially an issue in low-income countries (31). Moreover, the recently approved Rotavac and ROTASIIL vaccines require 3 doses, and may be used more frequently in low and middle income countries in the future (32). Even though overall efficacy is similar, observed effectiveness may be lower if the full series is not consistently administered on time. Additionally, to estimate the potential impact of rotavirus vaccination alone without any other interventions, we assume that the case fatality rate for rotavirus is fixed at its value from 2015 (3). However, rotavirus mortality has declined over time, even in locations that have not introduced rotavirus vaccination, which may be partially due to improved treatment, such as with oral rehydration solution (33). If so, we may have overestimated the proportional impact of rotavirus vaccination. We also assume that the probability of receiving one dose compared with two doses of vaccine are independent; however, in reality, a child who receives their first dose is more likely to get a second. Moreover, if children who receive a single dose of vaccine have lower socioeconomic status and poorer health outcomes than children who receive two doses (34), we might have overestimated the impact of a single dose of vaccine.

While other dynamic models of rotavirus vaccine introduction have been published, most have focused on a single country and/or a limited time horizon, making comparisons across countries and over time difficult (21). Other influential international models are static models, and cannot readily address the potential for indirect effects and the changing age distribution of cases as we have done here (3). Moreover, as most LMICs introduced vaccination somewhat later, only short-term empirical data are available. Models are needed to predict transmission dynamics in the long term. Incorporating information on the time since vaccine introduction allowed us to show that it might take several years to attain the full benefit of vaccination and that both doses are needed. This information is useful for cost-effectiveness analyses, as the impact of vaccination shortly after vaccine introduction might underestimate the overall impact once steady-state dynamics have been reached, due to gradually increasing coverage and indirect benefits (35). Our results highlight the high potential impact of rotavirus vaccination worldwide.

## Data Availability

All updated model code will be available on GitHub at the time of publication. Model output data files and input coverage estimates are proprietary of the Vaccine Impact ModelingConsortium and will not be publicly available. Validation data is the property of the Global Rotavirus Surveillance Network and will not be publicly available.

## Data sharing

All updated model code will be available on GitHub at the time of publication. Model output data files and input coverage estimates are proprietary of the Vaccine Impact Modeling Consortium and will not be publicly available.

## Acknowledgements

We would like to acknowledge contributions from the following GRSN contributors: Tishkova Faina, Bobokhonova Makhtob, Sulaimonova Sudoba, Mulladzhanova Manizha, Gafurov Oyatullo, Sodikova Umeda, Abdurakhmanova Salomat, Nazurdinov Anvar, Yunusova Midzhgona, Telahun Teka, Abebe Habtamu, Almaz Abebe, Frehiwot Kebede, Emmanuel Addo Yobo, Mawussi Godonou, Murithi Inoti, Kennedy Maore, Lilian Mbirithu, Bernard Gatinu, Monica Gitonga, Mercy Kinyua, Beth Maina, Thomas Gwiri, Grace Irimu, Nkolo mviena, Eric kemajou Grâce, Ngoya Roger, N. Gonah, H Mujuru and I Ticklay

