## Appendix for "Predicting the long-term impact of rotavirus vaccination in 112 countries from 2006-2034: a transmission modeling analysis"

#### Transmission model equations

The model is a system of ordinary differential equations:

$$\frac{dM_i}{dt} = (1 - c_1 v_{1i})(1 - cv_{2i})\delta M_{(i-1)} - M_i(\mu + \delta + \eta)$$

$$when\ i = 0\ then\ \delta M_{(i-1)} = number\ of\ births$$

$$\frac{dS_{1i}}{dt} = \omega R_i + \mu M_i + (1 - cv_{1i})(1 - cv_{2i})\delta S_{1(i-1)} - S_{1i}(\delta + \lambda_i + \eta)$$

$$\frac{dI_{ni}}{dt} = \lambda_i S_{ni} - \gamma I_{ni} - \delta I_{ni} + (1 - cv_{1i})(1 - cv_{2i})\delta I_{n(i-1)} - \eta I_{ni} \qquad (\text{For } n = 1..4)$$

$$\frac{dS_{ni}}{dt} = \alpha_{(n-1)}\gamma I_{(n-1)i} + (1 - cv_{1i})(1 - cv_{2i})\delta S_{n(i-1)} - S_{ni}(\delta + \lambda_i + \eta) \qquad (\text{For } n = 2..4)$$

$$\frac{dR_i}{dt} = \sum_{n=1}^{3}(1 - \alpha_n)\gamma I_{ni} + \gamma I_{4i} - \omega R_i - \delta R_i + (1 - cv_{1i})(1 - cv_{2i})\delta R_{(i-1)} - \eta R_i$$

$$\frac{dV_i}{dt} = \sum_{j=1}^{2}(cv_{ji})\left(\sum_{n=1}^{3}(1 - \alpha_n)\delta\big(M_{(i-1)} + S_{n(i-1)} + I_{n(i-1)}\big) + \delta\big(S_{4(i-1)} + I_{4(i-1)}\big)\right) + \delta V_{(i-1)} - V_i(\omega + \delta + \eta)$$

$$\frac{dS_{V1i}}{dt} = \omega(V_i + R_{Vi}) - \lambda_i S_{V1i} - \delta S_{V1i} + (1 - cv_{2i})\delta S_{V1(i-1)} - \eta S_{V1i}$$

$$when\ i = 2 - 3\ months\ old\ then\ \delta S_{V1(i-1)} = 0$$

$$\frac{dI_{V1i}}{dt} = \lambda_i S_{V1i} - \gamma I_{V1i} - \delta I_{1i} + (1 - cv_{2i})\delta I_{V1(i-1)} - \eta I_{V1i}$$

$$when\ i = 2 - 3\ months\ old\ then\ \delta I_{V1(i-1)} = 0$$

$$\frac{dS_{V2i}}{dt} = \alpha_1\gamma I_{V1i} + cv_{2i}\alpha_1\delta\big(M_{(i-1)} + S_{1(i-1)} + S_{V1(i-1)}\big) + (1 - cv_{2i})\delta S_{V2(i-1)} - S_{V2i}(\eta + \delta + \lambda_i)$$

$$\frac{dI_{Vni}}{dt} = \lambda_i S_{Vni} + cv_{2i}\alpha_n\delta\big(I_{n(i-1)} + I_{Vn(i-1)}\big) + (1 - cv_{2i})\delta I_{Vn(i-1)} - I_{Vni}(\gamma + \delta + \eta) \quad (\text{For } n = 2..4)$$

$$\frac{dS_{Vni}}{dt} = \alpha_{(n-1)}\gamma I_{V(n-1)i} + cv_{2i}\alpha_{(n-1)}\delta\big(S_{(n-1)(i-1)} + S_{V(n-1)(i-1)}\big) + (1 - cv_{2i})\delta S_{Vn(i-1)} - S_{Vni}(\eta + \delta + \lambda_i) \quad (\text{For } n = 3, 4)$$

$$\frac{dR_{Vi}}{dt} = \sum_{n=1}^{3}(1 - \alpha_n)\,\gamma I_{Vni} + \gamma I_{V4i} + (1 - cv_{2i})\delta R_{V(i-1)} - R_{Vi}(\eta + \delta + \omega)$$

where:

i = age group, defined as 0-1, 2-3 and 4-11 months of age, then one year age bands from 1-4 years, 5+ years.

$M_i$= those protected by maternal antibody in age group i

$S_{ni}$ = unvaccinated susceptibles to n[th] rotavirus infection (n =1 to 4) in age group i

$I_{ni}$ = unvaccinated infected by n[th] rotavirus infection (n=1 to 4) in age group i

$R_i$ = unvaccinated recovered and immune to rotavirus infection in age group i

$S_{Vni}$ = vaccinated susceptibles to n[th] rotavirus infection (n =1 to 4) in age group i

$I_{Vni}$ = vaccinated infected by n[th] rotavirus infection (n=1 to 4) in age group i

$R_{Vi}$ = vaccinated recovered and immune to rotavirus infection in age group I. These individuals do not gain immunity from vaccination but instead became immune through natural infection.

$V_i$ = those with long-term vaccine induced immunity in age group i

$\eta$ = all-cause mortality rate (set equal to 2015 country-specific birth rate)

$\mu$ = rate of loss of maternal immunity

$\delta$ = rate at which individuals in age group i age into age group (i+1)

$\gamma$ = rate of recovery from infection

$\omega$ = rate of loss of immunity

$c_i$ = the proportion who seroconvert to dose 1 or 2 of vaccine

$v_i$= the proportion receiving dose 1 or 2 of vaccine where vaccination is given at age 2 and 4 months.

$\alpha_n$ = risk of becoming re-susceptible after nth rotavirus infections

$\lambda_i$= force of infection; rate at which susceptible individuals become infected in age group i is expressed as:

$$\lambda_i(t) = \beta \sum_n \frac{I_{ni}(t)\xi_n}{N_i}$$

Where $\sum_n \frac{I_{nj}(t)\xi_n}{N_j}$ represents the proportion of symptomatic infectious individuals in age group i at n[th] infection at time $t$ ; $\xi_n$ is the proportion of $n$th infections that are symptomatic.

### Regression model details:

Predictor variables considered for inclusion in the regression model were: under 5 mortality rate, gross domestic product (GDP) per capita, total GDP, region, sub-region, birthrate, life expectancy and percent of the population living in a rural setting. Models with all possible subsets of variables were run. We assessed the performance of each model by comparing model fit statistics (AIC, BIC) and prediction performance using 80% of the full dataset as training data and 20% as test data. To assess prediction performance, we calculated correlation accuracy and mean absolute percent error (MAPE). We selected the model with the highest correlation accuracy and lowest MAPE.

The selected regression model is as follows:

$$Log(Y_j) = \beta_{1j}X_{1j} + \beta_{2j}X_{2j} + \beta_{3j}X_{3j} + \beta_{4j}X_{4j} + \beta_{5j}X_{5j} + \beta_{6j}X_{6j}$$

where:

$Y$ = mean age of severe rotavirus infection for each country based on GRSN data

$j$ = country

$\beta_1$= a continuous variable of the mortality rate per 1,000 for children >5 years old in 2015

$\beta_2$ = a continuous variable of the country-specific crude birth rate from 2015

$\beta_3$ = a dichotomous variable of the country-specific crude birth rate from 2015 $\begin{pmatrix} if < 20\ births\ per\ 1{,}000\ population & 0 \\ else & 1 \end{pmatrix}$

$\beta_4$ = a continuous variable of the country-specific life expectancy from 2015

$\beta_5$ = a continuous variable of the percent of the population living in rural settings

$\beta_6$= a continuous variable of the country-specific gross domestic product (gdp) per capita in 2015

### Transmission model parameters

***Table A1.*** *Input values, ranges and source for fixed parameters of the model*

| Parameter | | Value | Range | Source |
|---|---|---|---|---|
| **Not immune after:** | | | | |
| 1st infection | $\alpha_1$ | 0.61 | NA | (1) |
| 2nd infection | $\alpha_2$ | 0.48 | NA | (1) |
| 3rd infection | $\alpha_3$ | 0.33 | NA | (1) |

| | | | | |
|---|---|---|---|---|
| **Probability of symptoms:** | | | | |
| 1st infection | $\xi_1$ | 0.30 | NA | (1) |
| 2nd infection | $\xi_2$ | 0.28 | NA | (1) |
| 3rd infection | $\xi_3$ | 0.18 | NA | (1) |
| 4th infection | $\xi_4$ | 0.21 | NA | (1) |
| **Probability of severe disease:** | | | | (1) |
| 1st infection | $\chi_1$ | 0.17 | NA | (1) |
| 2nd infection | $\chi_2$ | 0.23 | NA | (1) |
| 3rd infection | $\chi_3$ | 0.24 | NA | (1) |
| 4th infection | $\chi_4$ | 0.18 | NA | (1) |
| **Relative infectiousness of non – 1° infections** | | 0.25 | NA | (1) |
| **Duration of:** | | | | |
| maternal immunity (days) | $\mu$ | 182 | NA | (2) |
| immunity (years) | $\omega$ | 250 | NA | Calibrated* |
| infection (days) | $\gamma$ | 7 | NA | (2) |
| **Vaccine Immunogenicity:** | | | | |
| Low-income country | c | 0.63 | (0.58, 0.67) | (3) |
| Middle-income country | c | 0.74 | (0.63, 0.86) | (3) |

* Duration of immunity was calibrated to value that ensured a high correlation between regression model and transmission model estimated mean age of infection. See Figure 2

### Regression model details for estimating age of infection

The dichotomous variable of crude birth rate was incorporated into the regression model after formal model selection to increase the correlation between the mean ages estimated by the linear regression to those estimated from the transmission model (Figure A1). The crude birth rates and life expectancy variables were informed by UNWPP data(4), while the variables for percent rural, GDP and under 5 mortality were informed by data from the World Bank. The country-specific mean ages of severe infection estimated by this regression model are in table A2.

***Figure A1.*** *Correlation of country-specific mean age of infection (weeks) estimated from a linear regression using GRSN surveillance data (y-axis) and a transmission model (x-axis). The 112 VIMC countries are color coded by World Bank income status.*

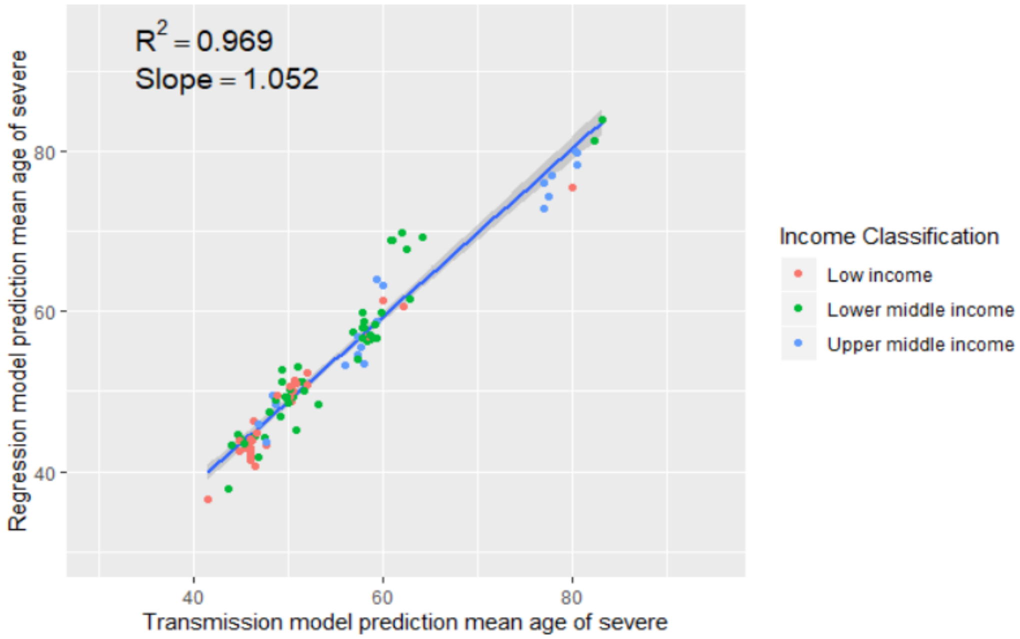


***Table A2.*** *Country-specific mean age of infection (95% confidence interval) estimated from regression model and calculated $R_0$ (range) used in transmission model. The estimates of $R_0$ are high as our calculation does not account for repeated infections. Rather than being interpreted as the true $R_0$ for a country, these values allow us to capture the transmission process in our model. Other models accounting for these features have estimated reproduction numbers that are much smaller (5,6). However, a recent model for Malawi estimated a reproductive number of 78.8 (7).*

| **Country** | **Estimated $R_0$ (Range)** | **Estimated Mean Age, weeks (95% Confidence Interval)** |
|---|---|---|
| **Afghanistan** | 71 (38, 92) | 47 (36, 86) |
| **Albania** | 55 (45, 113) | 74 (36, 90) |
| **Algeria** | 79 (46, 112) | 50 (35, 85) |
| **Angola** | 82 (35, 91) | 38 (34, 88) |
| **Armenia** | 53 (46, 114) | 73 (34, 85) |
| **Azerbaijan** | 58 (42, 118) | 65 (32, 90) |
| **Bangladesh** | 64 (42, 113) | 58 (33, 89) |
| **Belarus** | 54 (43, 110) | 71 (35, 90) |
| **Belize** | 75 (44, 119) | 51 (32, 88) |
| **Benin** | 70 (37, 98) | 45 (32, 84) |
| **Bhutan** | 52 (42, 95) | 71 (38, 88) |
| **Bolivia, Plurinational State of** | 68 (40, 104) | 54 (35, 91) |
| **Bosnia and Herzegovina** | 53 (46, 113) | 76 (35, 87) |

| | | |
|---|---|---|
| **Burkina Faso** | 71 (37, 91) | 44 (34, 85) |
| **Burundi** | 72 (37, 94) | 43 (33, 84) |
| **Cabo Verde** | 69 (46, 113) | 54 (33, 82) |
| **Cambodia** | 66 (42, 110) | 54 (33, 84) |
| **Cameroon** | 67 (34, 79) | 45 (38, 88) |
| **Central African Republic** | 60 (29, 73) | 44 (36, 90) |
| **Chad** | 70 (33, 94) | 40 (29, 85) |
| **China** | 57 (44, 113) | 70 (35, 89) |
| **Colombia** | 61 (45, 108) | 66 (37, 88) |
| **Comoros** | 69 (36, 101) | 48 (33, 92) |
| **Congo** | 72 (38, 101) | 45 (33, 86) |
| **Congo, the Democratic Republic of the** | 72 (34, 88) | 43 (35, 90) |
| **Cote d'Ivoire** | 66 (35, 90) | 44 (32, 84) |
| **Cuba** | 56 (45, 136) | 73 (30, 90) |
| **Djibouti** | 63 (40, 107) | 53 (31, 84) |
| **Ecuador** | 76 (46, 129) | 52 (31, 85) |
| **Egypt** | 74 (42, 101) | 50 (37, 88) |
| **El Salvador** | 54 (43, 107) | 70 (35, 88) |
| **Eritrea** | 70 (39, 98) | 48 (34, 87) |
| **Ethiopia** | 70 (39, 112) | 48 (30, 87) |
| **Fiji** | 68 (38, 106) | 51 (33, 91) |
| **Gambia** | 70 (35, 96) | 45 (33, 90) |
| **Georgia** | 52 (42, 123) | 73 (31, 89) |
| **Ghana** | 67 (37, 88) | 49 (37, 88) |
| **Guatemala** | 74 (42, 107) | 52 (35, 91) |
| **Guinea** | 69 (36, 85) | 45 (36, 86) |
| **Guinea-Bissau** | 65 (33, 91) | 45 (32, 90) |
| **Guyana** | 70 (42, 99) | 52 (36, 87) |
| **Haiti** | 62 (37, 108) | 53 (30, 89) |
| **Honduras** | 71 (43, 108) | 55 (36, 90) |
| **India** | 64 (41, 105) | 56 (34, 87) |
| **Indonesia** | 69 (42, 108) | 53 (34, 88) |
| **Iran, Islamic Republic of** | 59 (44, 115) | 66 (34, 90) |
| **Iraq** | 80 (40, 101) | 46 (36, 91) |
| **Jamaica** | 55 (46, 111) | 70 (35, 84) |
| **Jordan** | 74 (43, 127) | 52 (30, 90) |
| **Kenya** | 68 (39, 89) | 49 (38, 86) |
| **Kiribati** | 70 (40, 103) | 50 (34, 87) |
| **Korea, Democratic People's Republic of** | 54 (41, 110) | 69 (34, 90) |
| **Kosovo** | 50 (45, 112) | 78 (35, 88) |
| **Kyrgyzstan** | 69 (43, 96) | 53 (38, 86) |
| **Lao People's Democratic Republic** | 67 (42, 98) | 52 (35, 83) |

| | | |
|---|---|---|
| **Lesotho** | 56 (29, 77) | 48 (34, 91) |
| **Liberia** | 67 (37, 90) | 48 (36, 89) |
| **Macedonia, the former Yugoslav Republic of** | 53 (46, 106) | 74 (37, 85) |
| **Madagascar** | 69 (37, 92) | 49 (37, 91) |
| **Malawi** | 69 (37, 96) | 46 (34, 86) |
| **Mali** | 72 (34, 92) | 42 (32, 89) |
| **Marshall Islands** | 67 (41, 107) | 52 (33, 85) |
| **Mauritania** | 71 (40, 102) | 47 (32, 83) |
| **Micronesia, Federated States of** | 68 (42, 98) | 52 (36, 84) |
| **Moldova, Republic of** | 47 (42, 99) | 79 (37, 89) |
| **Mongolia** | 72 (41, 102) | 50 (35, 89) |
| **Morocco** | 72 (47, 112) | 55 (35, 85) |
| **Mozambique** | 67 (33, 84) | 45 (35, 89) |
| **Myanmar** | 47 (40, 102) | 72 (34, 86) |
| **Namibia** | 75 (36, 110) | 43 (29, 91) |
| **Nepal** | 64 (40, 102) | 56 (35, 91) |
| **Nicaragua** | 68 (45, 109) | 56 (35, 86) |
| **Niger** | 78 (35, 89) | 40 (35, 90) |
| **Nigeria** | 68 (31, 85) | 41 (33, 90) |
| **Pakistan** | 69 (39, 100) | 50 (35, 88) |
| **Palestine, State of** | 76 (43, 114) | 50 (33, 88) |
| **Papua New Guinea** | 67 (39, 107) | 49 (31, 85) |
| **Paraguay** | 72 (46, 107) | 53 (36, 83) |
| **Peru** | 75 (46, 105) | 52 (37, 87) |
| **Philippines** | 69 (42, 125) | 53 (29, 88) |
| **Rwanda** | 71 (39, 104) | 49 (34, 91) |
| **Samoa** | 75 (42, 108) | 50 (35, 90) |
| **Sao Tome and Principe** | 73 (41, 115) | 49 (31, 89) |
| **Senegal** | 73 (42, 101) | 47 (35, 82) |
| **Serbia** | 53 (44, 109) | 73 (36, 90) |
| **Sierra Leone** | 62 (32, 78) | 45 (35, 87) |
| **Solomon Islands** | 77 (46, 103) | 49 (36, 82) |
| **Somalia** | 72 (33, 81) | 41 (36, 88) |
| **South Africa** | 72 (38, 104) | 45 (31, 85) |
| **South Sudan** | 66 (33, 88) | 45 (34, 89) |
| **Sri Lanka** | 56 (46, 121) | 71 (33, 86) |
| **Sudan** | 72 (40, 92) | 47 (37, 85) |
| **Swaziland** | 64 (34, 92) | 45 (31, 85) |
| **Syrian Arab Republic** | 67 (41, 102) | 55 (36, 89) |
| **Tajikistan** | 73 (40, 113) | 50 (32, 91) |
| **Tanzania, United Republic of** | 71 (40, 94) | 46 (35, 83) |
| **Thailand** | 55 (44, 133) | 72 (30, 90) |

| | | |
|---|---|---|
| **Timor-Leste** | 70 (40, 107) | 51 (33, 88) |
| **Togo** | 66 (37, 84) | 47 (37, 84) |
| **Tonga** | 73 (43, 106) | 50 (35, 85) |
| **Tunisia** | 57 (45, 109) | 70 (36, 88) |
| **Turkmenistan** | 79 (42, 111) | 45 (32, 84) |
| **Tuvalu** | 72 (40, 102) | 51 (36, 91) |
| **Uganda** | 73 (38, 90) | 44 (35, 84) |
| **Ukraine** | 47 (44, 114) | 79 (33, 85) |
| **Uzbekistan** | 68 (44, 107) | 54 (34, 84) |
| **Vanuatu** | 75 (44, 114) | 48 (32, 83) |
| **Venezuela, Bolivarian Republic of** | 90 (42, 106) | 42 (36, 91) |
| **Viet Nam** | 52 (47, 115) | 75 (34, 83) |
| **Yemen** | 70 (39, 97) | 49 (35, 88) |
| **Zambia** | 72 (39, 97) | 44 (33, 82) |
| **Zimbabwe** | 66 (35, 86) | 47 (36, 88) |

#### Calculating deaths and DALYs averted

To calculate total deaths, we estimated the number of severe rotavirus cases for each country and year based on the number of previous rotavirus infections (with first infections being most severe and subsequent infections being less likely to cause severe disease) and then multiplied this quantity by the estimated rotavirus case fatality risk for each country and age group, based on data from the Global Burden of Disease Study(8). The case fatality risk was based on the ratio of the total number of expected deaths for each country in the Global Burden of Disease study prior to 2005 (before rotavirus vaccination became available) divided by the modeled number of severe cases over the same period. We also estimated the impact of rotavirus vaccination on DALYs averted, combining information on the number of deaths and severe cases averted as well as life expectancy.

#### Validation of model predictions using surveillance data

Countries selected for inclusion in the model validation exercise were those with at least 3 years of surveillance data prior to and after the year of vaccine introduction and an average of at least 20 rotavirus-positive cases per year over the study period (to allow for reliable calculation of confidence intervals). For each of the 10 countries included in the analysis, we used all years with available GRSN data prior to vaccine introduction to calculate the average proportion of GE hospital admissions positive for rotavirus prior to vaccine introduction. We then compared this pre-vaccine average with the proportion of hospital admissions positive for rotavirus for each year after vaccine introduction, excluding the year vaccination was introduced. The percent reduction in rotavirus positivity for each post-vaccine year was calculated by taking the ratio of these two proportions. We then compared the yearly percent reduction in rotavirus positivity from the GRSN data with our modeled percent reduction in severe rotavirus cases on a yearly basis. The overall data available for the 10 countries used in the validation analysis is shown in **Table A3**. The relative percent reductions in rotavirus positivity were conceptualized as approximating a risk ratio, and relevant methods were used to calculate the 95% confidence interval for each year.

As a sensitivity analysis, we also compared our model with the percent reduction in rotavirus positive GE admissions. To do this, we first calculated the average yearly number of rotavirus positive hospital admissions prior to vaccine introduction. We then compared this pre-vaccine average number of rotavirus positive admissions to the number of rotavirus positive admissions in each post-vaccine year as a proportion to estimate the yearly percent reduction in rotavirus admissions. Similar to the analysis calculating percent reduction in rotavirus positivity described above and shown in Table A3, we calculated confidence intervals using a risk ratio. The resulting plots are shown in Figures A5 (under 1) and A6 (under 5).

**Table A3.** Total number of rotavirus + and rotavirus – admissions for all children 0-59 months for the 10 countries included in the validation analysis.

| **Country** | **Pre-vaccine introduction** | | | **Post-vaccine introduction** | | |
|---|---|---|---|---|---|---|
| | **Years** | **Total Rota + Admissions (annual range)** | **Total Rota- Admissions (annual range)** | **Years** | **Total Rota + Admissions (annual range)** | **Total Rota - Admissions (annual range)** |
| Cameroon | 6 | 1278 (33-456) | 1920 (123-610) | 4 | 458 (55-188) | 1712 (231-679) |
| Ethiopia | 6 | 515 (45-114) | 1769 (136-419) | 5 | 213 (25-56) | 880 (144-244) |
| Fiji | 5 | 469 (38-132) | 636 (92-175) | 6 | 91 (2-38) | 469 (17-119) |
| Ghana | 3 | 1186 (274-517) | 953 (1-493) | 7 | 520 (4-131) | 1420 (24-293) |
| Kenya | 6 | 1251 (57-386) | 2236 (79-713) | 5 | 230 (2-110) | 959 (10-351) |
| Tajikistan | 3 | 1395 (179-694) | 2124 (438-965) | 4 | 1068 (165-375) | 3221 (680-978) |
| Tanzania | 4 | 453 (70-212) | 672 (108-290) | 6 | 1949 (27-839) | 7033 (169-2068) |
| Togo | 5 | 531 (74-140) | 363 (0-114) | 5 | 154 (4-60) | 318 (27-116) |
| Zambia | 6 | 2328 (208-502) | 3634 (369-712) | 5 | 1403 (1-464) | 3007 (127-1021) |
| Zimbabwe | 6 | 2944 (125-960) | 3297 (204-1008) | 5 | 1393 (6-492) | 4623 (70-1718) |

**Figure A2.** Predicted vaccine impacts on annual severe rotavirus cases for children under 1 year of age (model) and percent reduction in rotavirus hospital admissions (GRSN data) (y-axis) by number of years since vaccine introduction (x-axis) for 10 countries (shown in different panels). Modeled impacts are shown in solid lines, with the grey ribbon showing 95% simulation intervals. Impacts based on surveillance data are shown with points, with error bars to show uncertainty (error bars were calculated using standard methods, assuming that the percent reduction in positivity approximated a risk ratio).

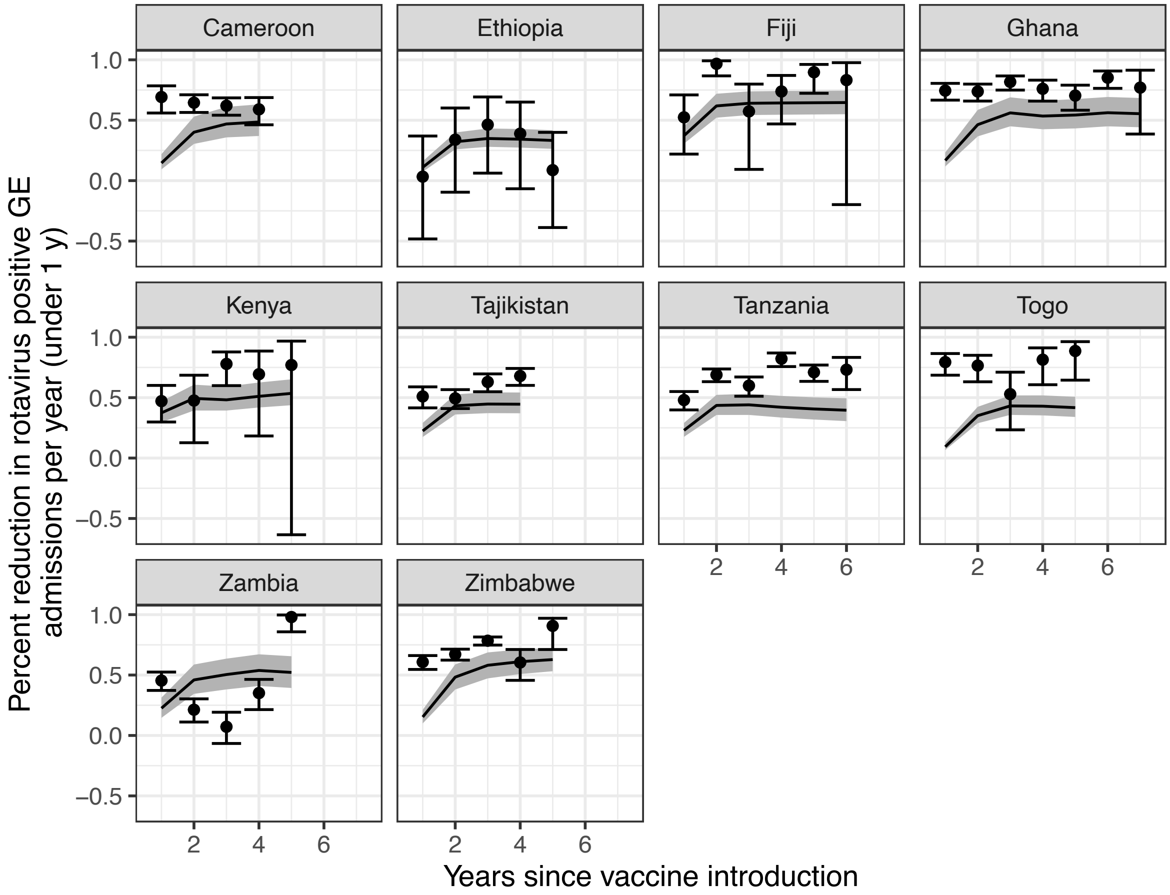


**Figure A3.** Predicted vaccine impacts on annual severe rotavirus cases for children under 5 years of age (model) and percent reduction in rotavirus hospital admissions (GRSN data) (y-axis) by number of years since vaccine introduction (x-axis) for 10 countries (shown in different panels). Modeled impacts are shown in solid lines, with the grey ribbon showing 95% simulation intervals. Impacts based on surveillance data are shown with points, with error bars to show uncertainty (error bars were calculated using standard methods, assuming that the percent reduction in positivity approximated a risk ratio).

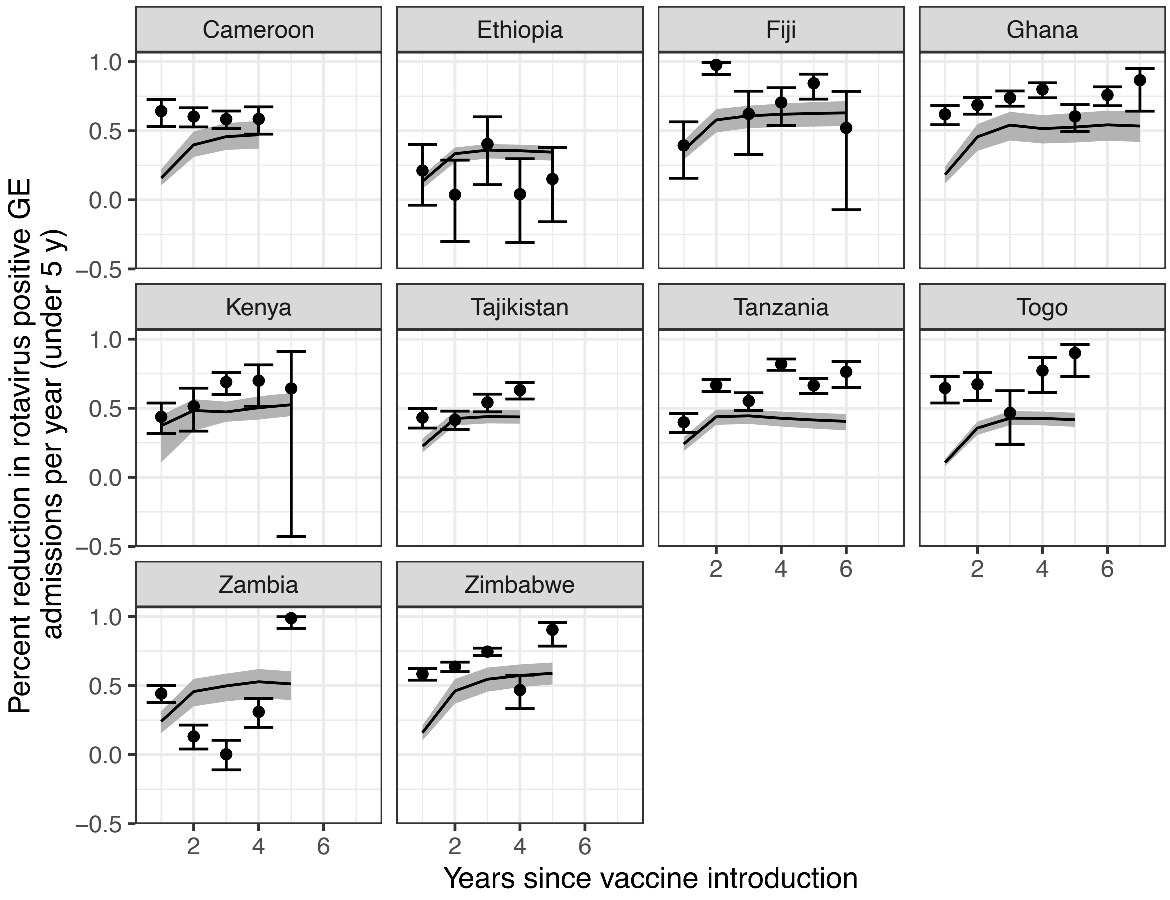


**Projected DALYs averted by country income level and world health region**

Low-income countries briefly outpace low/middle income countries shortly after rollout in terms of DALYs averted due to a few low-income countries in sub-Saharan Africa.

**Figure A4.** DALYs averted by country income level. Solid lines and circle shapes show impact under the default scenario and dashed lines and open squares show predicted impact under the best-case scenario.

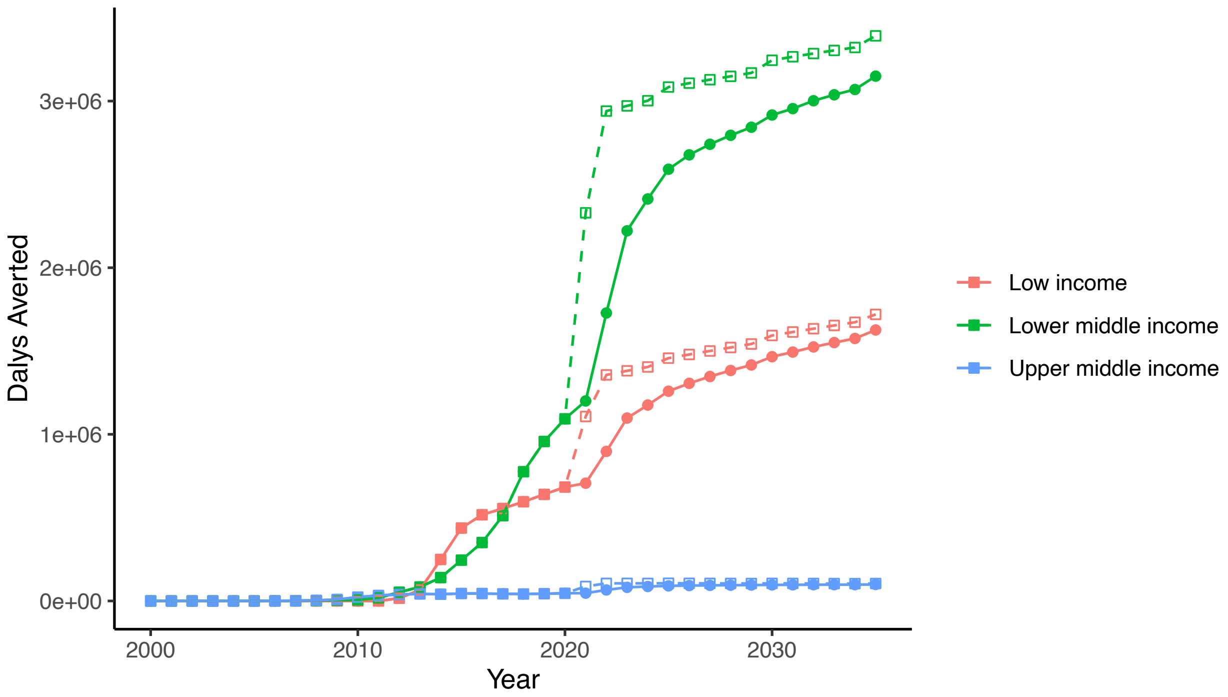


**Figure A5.** Predicted DALYs averted WHO region. Solid lines and open circles show impact under the best case scenario and dashed lines with solid circles show predicted impact under the best-case scenario

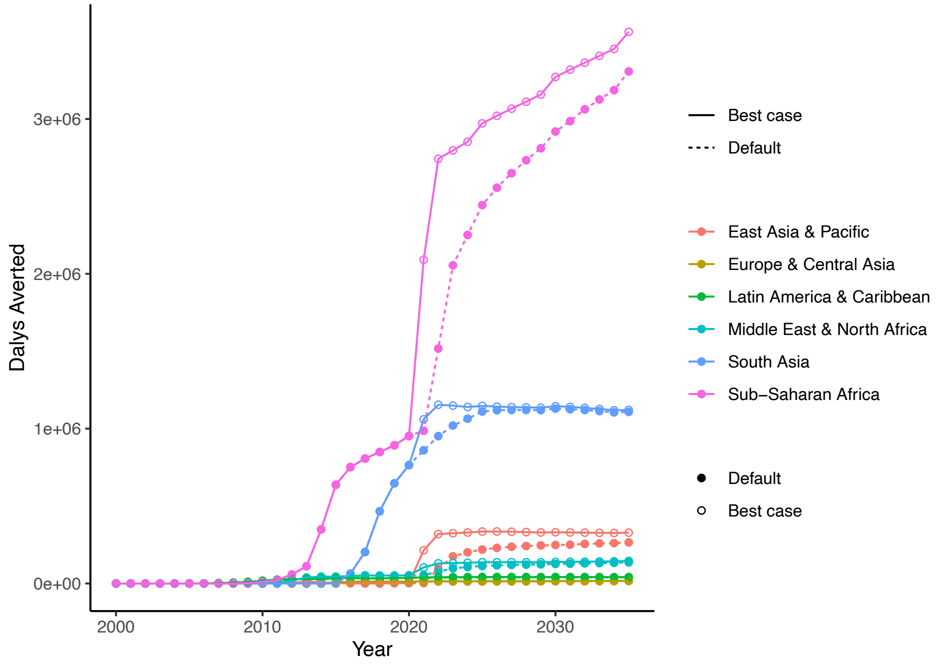


### Age-specific impacts on deaths

**Figure A6.** Deaths by age group over time, stratified by region.

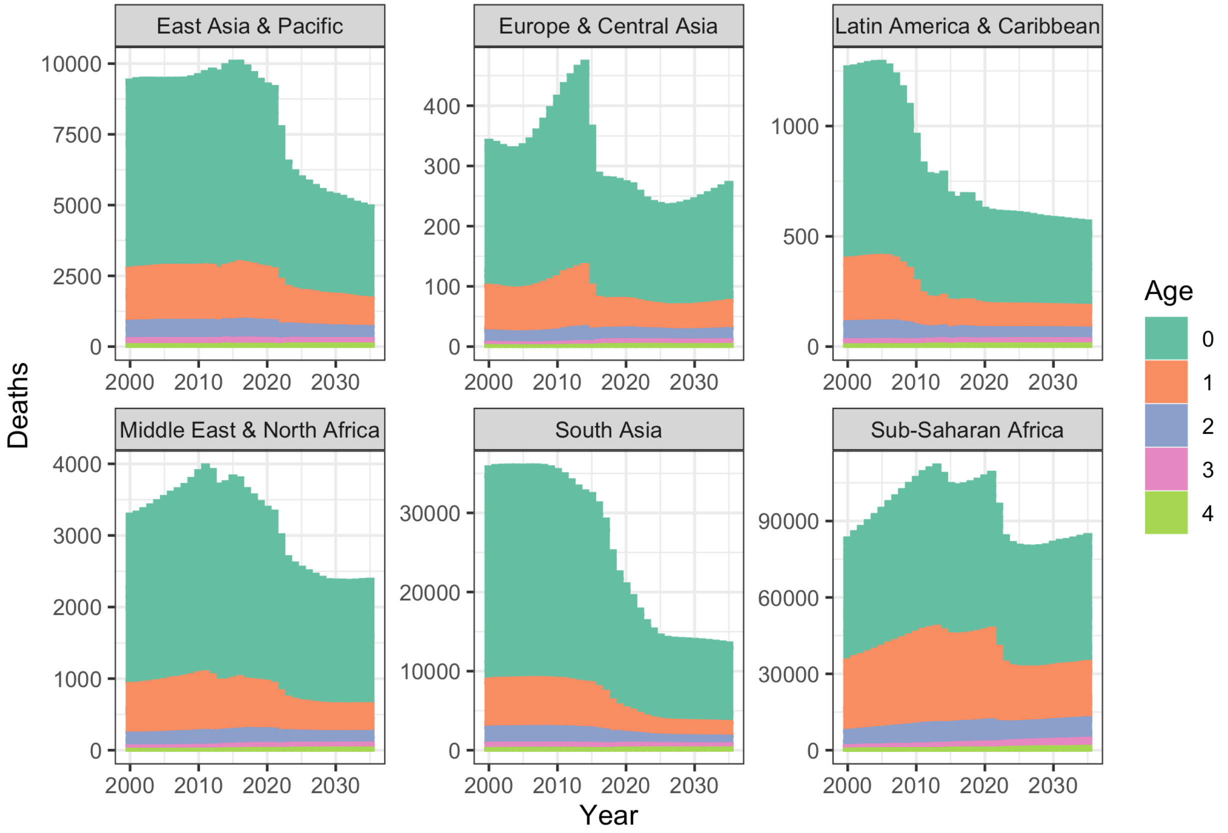
